# Self-perceived competence and its determinants among pre-intern (PRINT) doctors in Uganda

**DOI:** 10.1101/2024.01.12.24301259

**Authors:** Nelson Ssewante, Godfrey Wekha, Racheal Nalunkuma, Lawrence Katumba Sentongo, Bereta Sanyu, Moureen Namusoke, Ayub Nkwanga, Rachel Ahabwe, Vanessa Nalwoga Nantagya, Sharon Esther Kalembe, Catherine Nampeera, Phillip Musoke, Pauline Byakika-Kibwika

## Abstract

**Introduction:** Uganda still survives way below the recommended doctor-patient ratio. This problem could be solved by increasing the number of doctors produced in the country annually. Attempts at this are ongoing, however, this is challenged by the absence of quality assurance programs amidst lack of a universal medical curriculum. With inadequate supervision and limited resources in health facilities, transitioning from students’ life to clinical practice is perhaps the most challenging time for an intern doctor. We used the domains of competencies provided by the General Medical Council to assess levels of self-perceived competence and its determinants among pre-intern doctors (PRINTs) in Uganda.

**Methods:** An online cross-sectional study was conducted using a quantitative questionnaire distributed to confirmed pre-interns in Uganda. Self-perceived competence was determined by 4 Likert scale questions and participants were dichotomized into competent and incompetent using a standardized Bloom’s cutoff criterion. Determinants of self-perceived competence were determined by the ANOVA test.

**Results:** We obtained 142 entries. Of these, 68.3% were males; median age was 26 years (range: 22-49). Majority of the participants (78.9%) had no prior medical-related training and only a third (34.5%) had previous work experience. Overall scores were poor and very few participants were confident they attained a satisfactory level of competence through their medical training. Participants reported more competence in basic procedural skills (Mean score: 19.8±3.2/21), followed by Knowledge (Mean score:12.4±3.1/15), with surgical (Mean score:11.5±3.5/18) and Emergency skills (Mean score: 12.5±4.4/21) scoring least. Participants with previous work experience were more likely to report higher competence scores than their counterparts (91.7 vs 84.9, p=0.039).

**Conclusion:** The study shows that majority of PRINTs feel deficient in knowledge and skills to start clinical practice, with less competence in surgical skills and emergency care.

There is need to review the training curricula to ensure adequate clinical exposure experiences for a smooth transition to clinical practice.

## Background

According to the WHO, a doctor-to-patient ratio of 1: 1,000 is a necessary prerequisite in the bid to achieve optimal health care delivery to the population. Up to date, Uganda stands at 1:25,000 which puts the goal far beyond reach(1). This is compounded by a poor recruitment system leaving almost 50% of qualified doctors not employed by the government(2,3) and many migrating to foreign countries for job opportunities,leaving inexperienced intern doctors as major service providers in Uganda’s health system. Every year, over 500 doctors complete medical training with Bachelor of Medicine and Bachelor of Surgery (MBChB) from the different medical schools in Uganda (2). These institutions operate on different curricula to train medical students into prospective doctors. To ensure all junior doctors have certain acceptable practicing standards, Uganda has a policy that requires these fresh graduates to undergo a mandatory one-year internship program in a pre-determined hospital under the supervision of a senior doctor before the award of a permanent practicing license. However, because of the diversity in resource allocations in the different hospitals in the country, some intern doctors may be at a disadvantage and forced into a life of improvisation. This predisposes them to incompetent and error-prone practice an example being the famous case of Jinja Regional Referral Hospital where an unsupervised intern doctor allegedly cut the arm of a baby during a Caesarean section(4).

Over the years, the number of medical schools has increased as well as the number of graduating doctors (2,5). This is a positive record, especially intending to narrow the doctor-patient ratio and health system strengthening(6). However, the increase in number of universities comes with concerns of the quality of the doctors produced and for a long, this has been an unanswered question(5). This is especially due to; 1) partial privatization of education with inadequate quality assurance systems to ensure private institutions adhere to a particular set standard, 2) lack of a universal curriculum ‒ some universities for example run 5-year-while others a 5.5-year-long MBChB degree program, and 3) lack of a National Examination Program to assess the quality of qualifying doctors. As a result, the quality of fresh doctors is entirely in the hands of the training institutions save for the efforts of the trainee.

According to the General Medical Council (GMC), assessment of competence among trainee doctors is based on four domains: Domain 1 focuses on Knowledge, Skills, and Performance; Domain 2 Safety and Quality; Domain 3 Communication, Partnership, and Teamwork; Domain 4 Maintaining Trust(7). The council recommends that the first 3 competencies which cover the principles of history taking, clinical examination and therapeutics, and safe prescribing be emphasized during the fundamental training of health professionals. These are further reinforced together with the fourth competence during further training such as, internship and specialized training. With the diversity in training approaches at the different medical schools in the country, we set out to assess the levels of self-perceived competence and its determinants among pre-intern (PRINT) doctors based on the first domain (knowledge, skills, and performance).

## Methods

### Study design

This was an online cross-sectional study collecting quantitative data from PRINTs in Uganda between February and March 2022.

### Study population

We included PRINTs at the eight (8) medical schools in Uganda. A PRINT was defined as any student who had completed the final semester of their medical training in Bachelor of Medicine and Bachelor of Surgery at one of the recognized medical schools in Uganda. The public universities with medical schools in Uganda include Makerere University (MAK), Mbarara University of Science and Technology (MUST), Kabale University (KU), Gulu University (GU), and Busitema University (BU). Private universities include King Caesar International University, Kampala International University (KIU), and Islamic University in Uganda (IUIU).

### Sample size estimation and sampling procedure

The sample size was calculated using methods for cross-sectional studies by feeding the population size and estimated prevalence at a 95% confidence interval into an online sample size calculator(8). Considering an average of 500 doctors produced annually in Uganda, an estimated sample size of 218 participants was obtained.

Using this estimate, 218 potential participants were selected from the respective participating universities using a stratified random sampling approach. To ensure participants were randomly selected, we identified class leaders from each participating university (stratum) who provided lists of names for pre-interns including their contact details. Each name on the list was assigned a unique number and these were used to randomly select participants. Selected participants were contacted using direct phone calls, WhatsApp messages, and where applicable email messages to seek informed consent to participate.

### Selection criteria

A potential participant was eligible for this study upon informed consent if they had completed the MBChB course awaiting the Internship program. Participants who were ill, or unable to set up a good internet connection were automatically excluded.

### Data collection methods

This was an online study conducted using an electronic questionnaire designed with Google forms. The link to this Google form was shared with potential participants using their email addresses, and social media platforms particularly, WhatsApp Messenger.

The questionnaire was designed to comprise two sections:

Section I (Participant’s Information): Covering the demographic characteristics of participants which served as the independent variables in statistical analysis.

Section II (Competence assessment): This assessed the self-perceived levels of competence as regards knowledge and procedural skills using the 4-Likert scale. Knowledge was assessed using; “Not at all competent”, “Inadequately competent”, “Fairy competent” and “Adequately competent”. Procedural skills were classified into Basic skills, Medical skills (catering to both Internal Medicine and Pediatrics), Surgical skills, Obstetrics and Gynecology (OBGY) skills, and emergency medicine skills. These were assessed using; “Never performed nor observed”, “Never performed but observed”, “Can perform under supervision” and “Can perform without supervision”.

This questionnaire was developed in reference to the two previous studies(9,10) and the undergraduate clinical assessment logbooks.

### Quality control

The questionnaire was designed with appropriate skip logics and all questions were mandatory to avoid missing information. Pretesting of the questionnaires was done before data collection to assess for ambiguity and clarity of the questionnaire among 10 PRINTs whose entries were not part of the final dataset. The internal consistency of the scale used to assess self-perceived competency was calculated and a Cronbach’s alpha of 0.955 was obtained.

### Ethical approval and consent to participate

The study was conducted in accordance with the Declaration of Helsinki; approved by the Mulago Hospital Research and Ethics Committee under reference number MHREC 2201. Verbal informed consent was obtained from all participants before a link to the online questionnaire was shared. Additionally, the consent form was provided on the cover page of the Google Form and participants were asked to affirm whether they were willing to continue using prompts “Yes” or “No”. Those who chose “Yes” were linked to the questionnaire while those who chose “No” were not granted access to the questionnaire.

### Data analysis

Entries from the Google forms were downloaded and the dataset was imported into Microsoft Excel 2019 for cleaning and coding. The categorical variables were summarized as frequencies and percentages and numerical variables into means and standard deviations. To assess the level of self-perceived competence, the Likert scale was scored as; Adequately competent/ Can perform without supervision: 3, Fairly competent/Can perform under supervision: 2, Inadequately competent/Never performed but observed: 1, Not at all competent/Never performed nor observed: 0. The sub-total scores for each of the skill sub-groups were computed. Overall total scores were calculated by adding these subtotals and their percentages computed. Participants were dichotomized into competent and incompetent subgroups using an 80% score as the cutoff based on Bloom’s cutoff criterion. Total scores were used to determine the correlation with independent variables assessed using a one-way Analysis of Variance (ANOVA) testing model with a p<0.05 considered statistically significant.

### Results

A total of 142 participants (response rate 65.1%) were obtained. Of these, 68.3% were males with a median age of 26 years (IQR: 22-49). Majority of participants (78.9%) were direct entrants with no prior medical-related training and only a third (34.5%) had previous work experience (**Table 1**). About a third (35.2%) of participants trained from Makerere University; the rest trained from other participating institutions as shown in **Figure 1**.

**Table 1:** Characteristics of participants

| Characteristics | Frequency | Percentage |
| --- | --- | --- |
| **Age, [Median (IQR)]** | 26 (22-49) years |  |
| ≤26 years | 93 | 65.5 |
| >26 years | 49 | 34.5 |
| **Gender** |  |  |
| Male | 97 | 68.3 |
| Female | 45 | 31.7 |
| **Institution type** |  |  |
| Public | 108 | 76.1 |
| Private | 34 | 23.9 |
| **Had previous medical training** |  |  |
| No | 112 | 78.9 |
| Yes | 30 | 21.1 |
| **Previous medical training accomplished** |  |  |
| Diploma | 19 | 65.5 |
| Bachelor | 9 | 31.0 |
| Certificate | 1 | 3.5 |
| **Previous working experience** |  |  |
| None | 93 | 65.5 |
| Yes | 49 | 34.5 |
| **Duration of employment** |  |  |
| 1 to 2 years | 29 | 59.2 |
| 3 or more | 20 | 40.8 |
| **Facility level** |  |  |
| Health centre/clinic | 31 | 63.3 |
| Hospital | 16 | 32.7 |
| Pharmacy | 2 | 4.1 |

**Figure 1:**
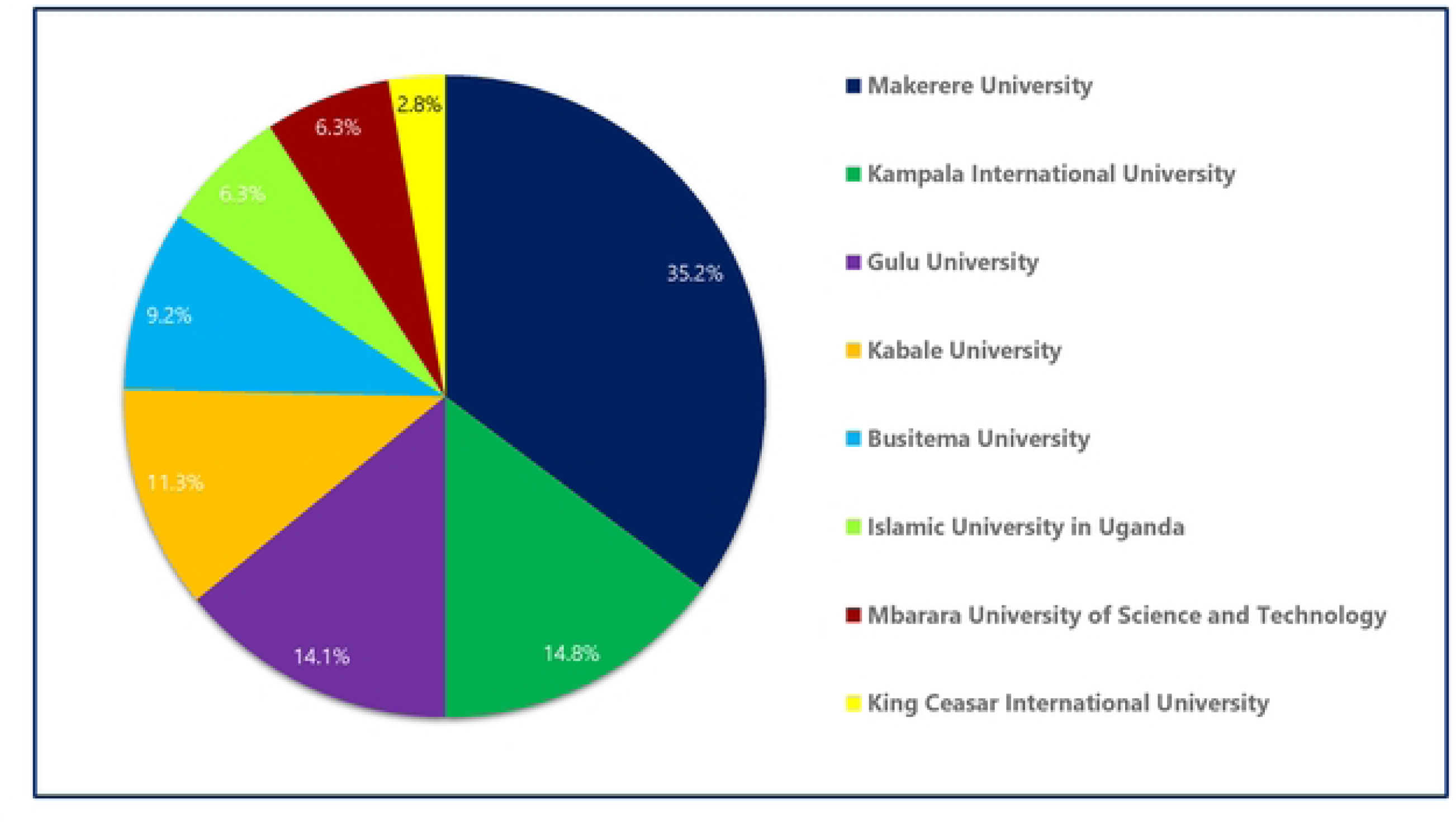
Participating Universities

From the participants’ perspective, majority were competent in history taking, physical examination, and formulation of differential diagnoses. However, they were skeptical about the interpretation of investigations and formulation of management plans (**Table 2**).

**Table 2:** Self-reported competence in knowledge skills

| Skill | Adequately competent | Fairly Competent | Inadequately Competent | Not at all competent |
| --- | --- | --- | --- | --- |
| History taking | 114(80.3) | 17(12) | 9(6.3) | 2(1.4) |
| Physical examination | 93(65.5) | 36(25.4) | 10(7) | 3(2.1) |
| Interpretation of investigations | 66(46.5) | 60(42.3) | 12(8.5) | 4(2.8) |
| Formulation of a logical differential diagnoses | 80(56.3) | 50(35.2) | 10(7) | 2(1.4) |
| Formulation of the logical management plan | 68(47.9) | 57(40.1) | 14(9.9) | 3(2.1) |

Overall, the scores were poor and very few participants were confident that they attained a satisfactory level of competence through their medical training (**Figure 2**).

**Figure 2:**
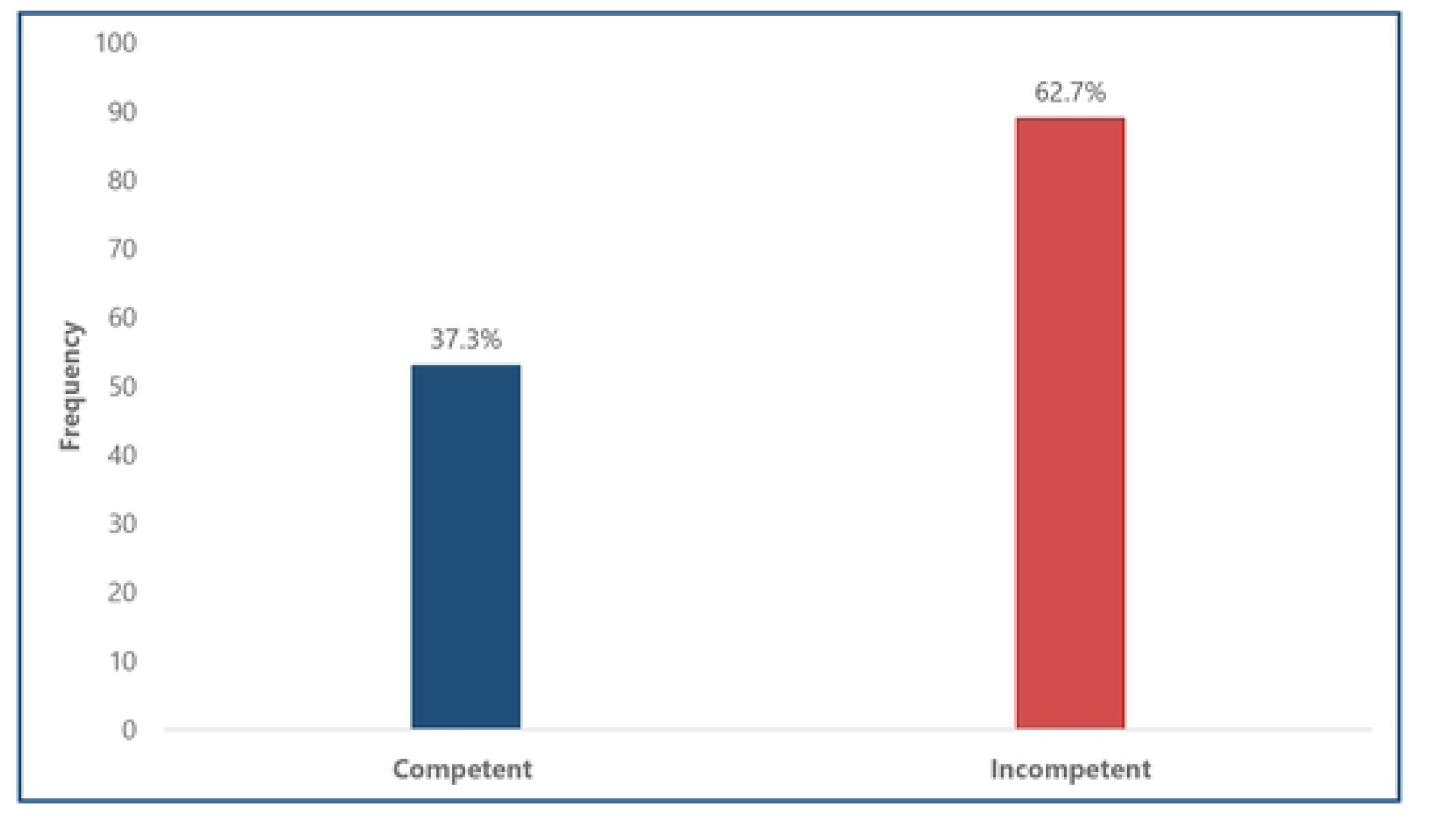
Overall Self-Perceived Competence among PRINTs

Participants reported more competence in general procedural skills (Mean score: 19.8±3.2/21), followed by knowledge skills (Mean score:12.4±3.1/15), with surgical (Mean score:11.5±3.5/18) and emergency skills (Mean score: 12.5±4.4/21) scoring least as shown in figure 3.

**Figure 3:**
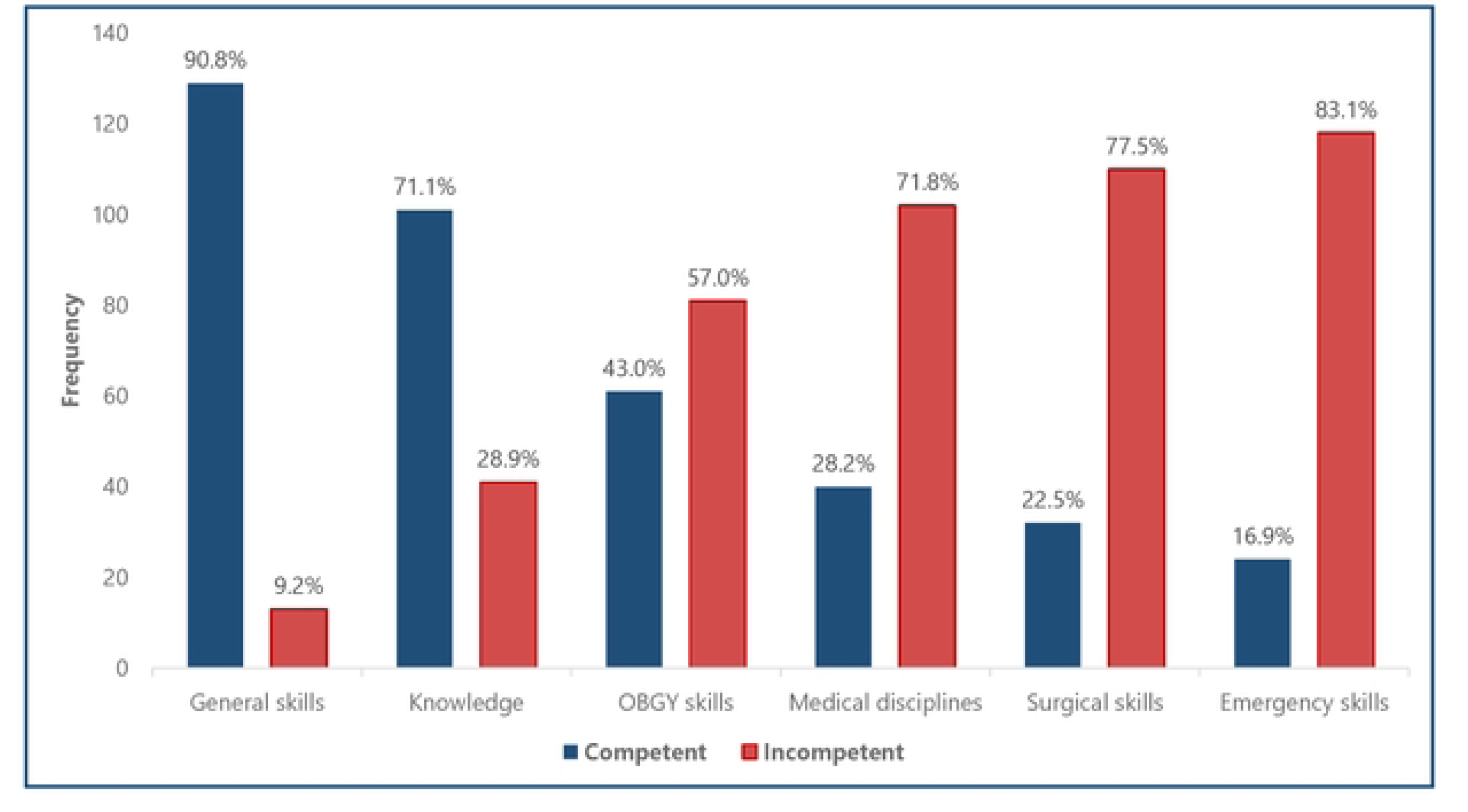
Self-perceived Competence by discipline

Among the general skills, nearly all believed were competent in setting up intravenous (IV) drips (95.1%) and cannulation (94.4%) with nasogastric (NG) tube insertion as the less-scored skill.

Childhood procedures were found to be more challenging. Most participants felt like they still needed supervision in the majority of the surgical skills.

Participants were less confident about lifesaving skills such as twin delivery, breech delivery, and C-section although they had observed them and majority believed they could perform such procedures with adequate supervision.

In the emergency management skills, participants generally reported low levels of competency except for Glasgow Coma Scale (GCS) monitoring (**Table 3**).

**Table 3:**
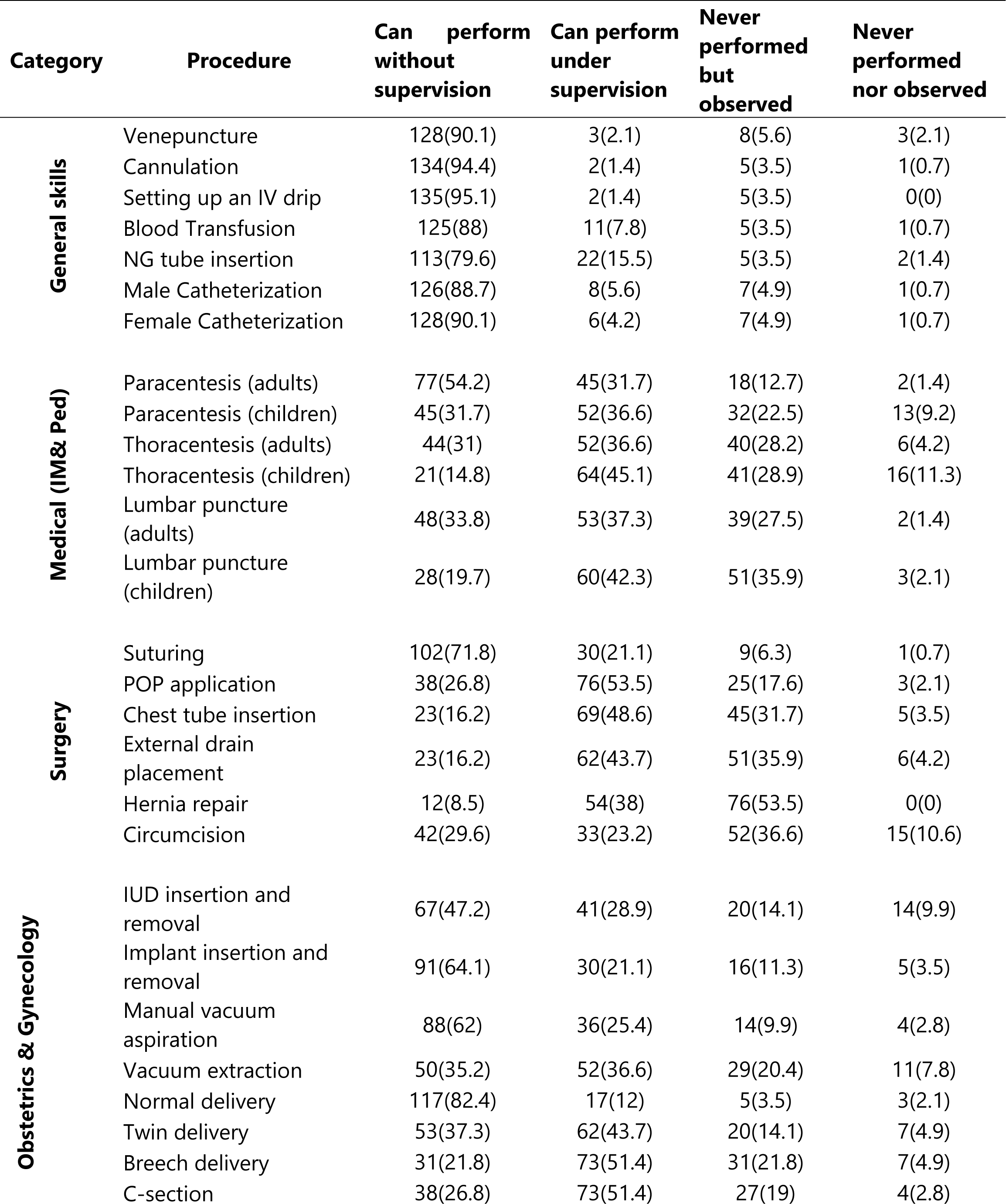
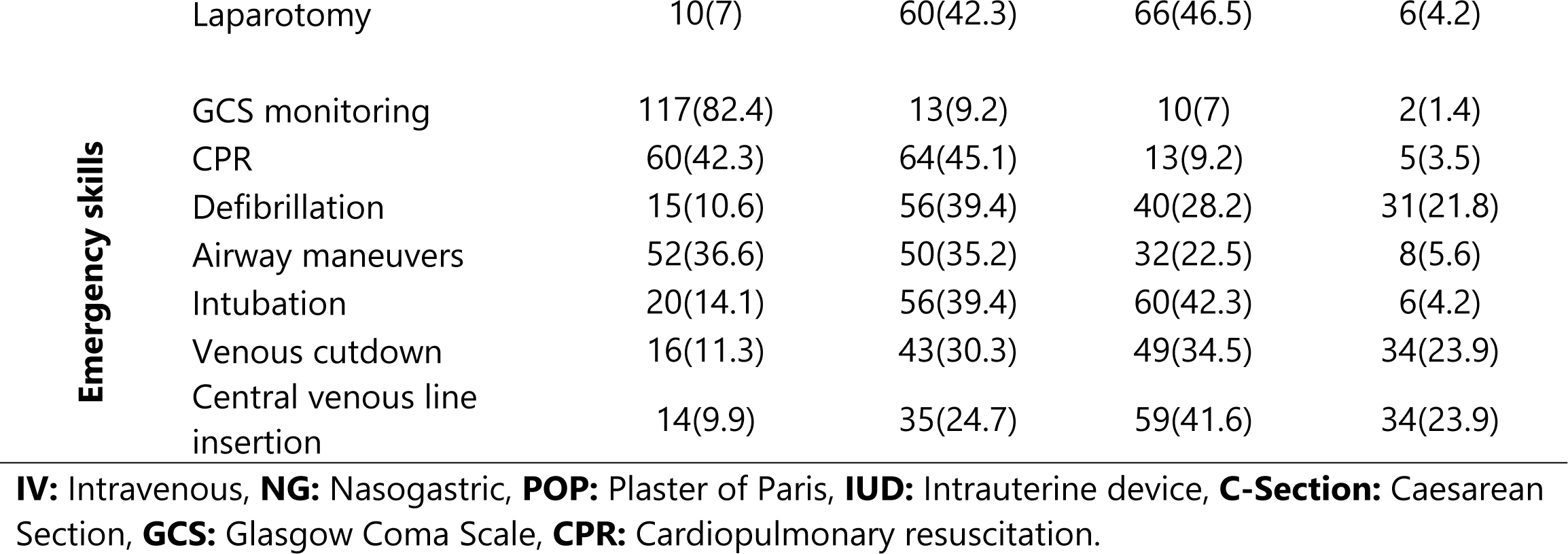
Self-reported competence in procedural skills.

### Determinants of self-perceived competence among participants

After a One-way ANOVA, participants with previous work experience in a clinical setting were more likely to report higher competency scores than their counterparts (91.7 vs 84.9) p=0.039 (**table 4**). The variations in median scores among those with and those without previous work experience are shown in figure 4.

**Table 4:** Determinants of self-perceived competency

| **Factor** | **Mean** | **Standard deviation** | **p-value** |
| --- | --- | --- | --- |
| **Age** |  |  | 0.639 |
| >25 years | 88.0 | 19.8 |  |
| ≤25 years | 86.5 | 17.4 |  |
| **Gender** |  |  | 0.087 |
| Female | 83.3 | 16.0 |  |
| Male | 89.1 | 19.6 |  |
| **Institution** |  |  | 0.479 |
| Private | 89.3 | 19.6 |  |
| Public | 86.6 | 18.5 |  |
| **Previous medical-related training** |  |  | 0.320 |
| No | 86.5 | 19.3 |  |
| Yes | 90.3 | 16.0 |  |
| **Previous training level** |  |  | 0.330 |
| Bachelor | 83.2 | 14.5 |  |
| Certificate | 88.0 | 0.0 |  |
| Diploma | 93.0 | 16.5 |  |
| **Previous working experience** |  |  | 0.039 |
| None | 84.9 | 18.2 |  |
| Yes | 91.7 | 19.1 |  |
| **Employing Facility** |  |  | 0.743 |
| Health Centre/clinics | 93.4 | 14.6 |  |
| Hospital | 89.1 | 25.6 |  |
| Pharmacy | 88.0 | 31.1 |  |
| **Duration of employment** |  |  | 0.733 |
| 1-2 years | 92.5 | 20.4 |  |
| 3 or more years | 90.6 | 17.4 |  |

**Figure 4:**
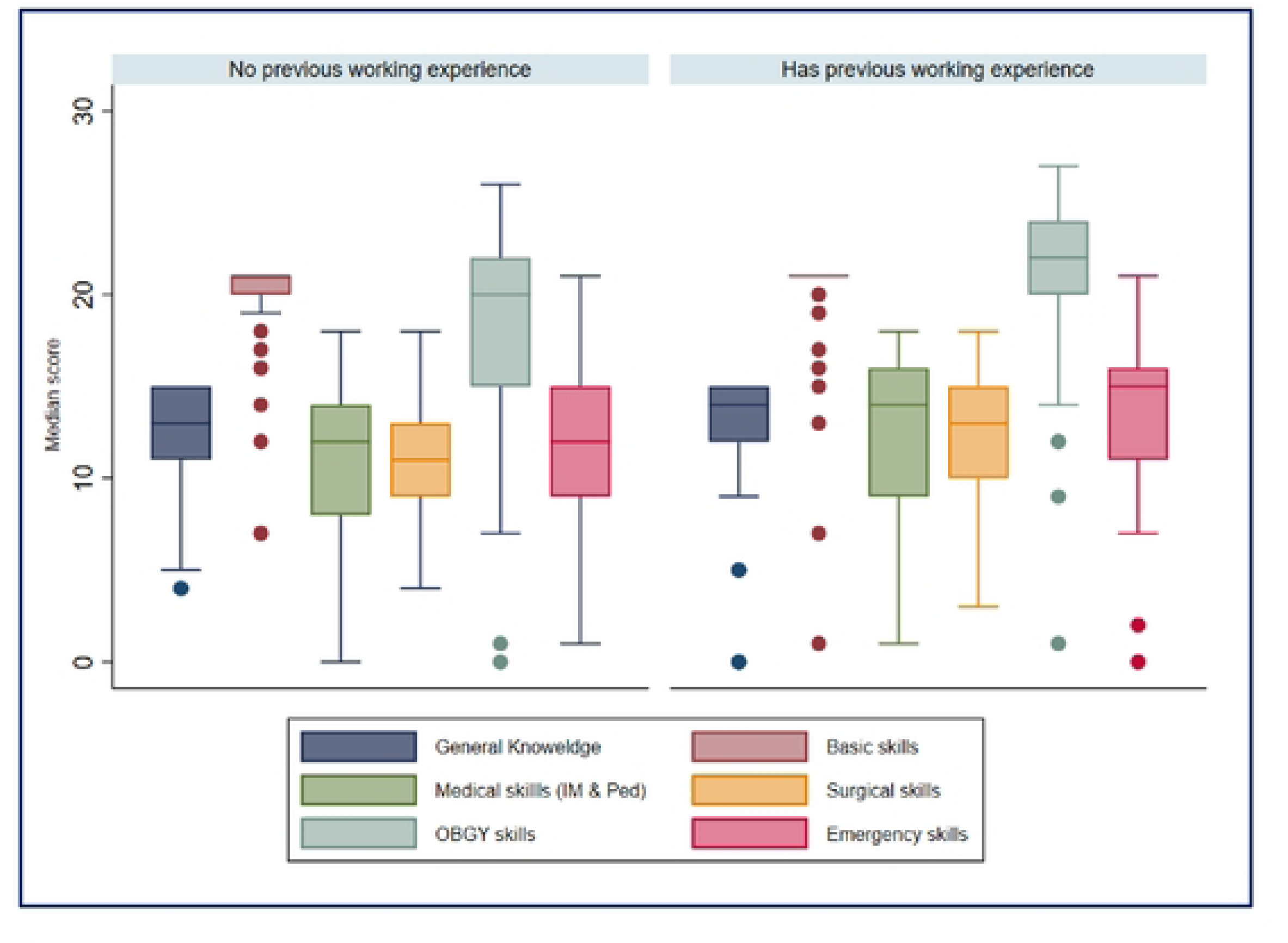
Median score differences based on previous working experience

## Discussion

This study reports on self-perceived competence among PRINTs and its determinants. Findings demonstrate a good number of PRINTs were confident about their level of competence in knowledge, and general procedural skills but less competent in other procedural skills in the major medical disciplines. Previous work experience in a clinical setting was the only statistically significant factor associated with self-reported competence.

Training a student into a medical doctor takes more than instilling knowledge. It should impart confidence to translate the acquired knowledge into practice. In this study, nearly all participants believed they had gained enough knowledge to clerk patients to the gist of the problem. However, some still felt less competent in translating information obtained from their clerkship to formulate logical differential diagnoses and management plans for their patients. This is probably influenced by low levels of self-perceived competence in the interpretation of investigations. Literature shows that investigations provide a significant challenge to inexperienced/junior doctors which limits their ability to make conclusive or even correct diagnoses(11,12). Although virtually all patients presenting for hospital care have at least one investigation requested by an attending doctor, most investigations present a significant challenge during interpretation. Errors in interpretation of investigations results in significant adverse effects including misdiagnosis, overspending on additional and often unnecessary investigations, and poor prognosis owed to mismanagement(13). This finding, therefore, could mean that although the PRINTs may be competent in clerkship, translation to practice and patient care may prove a key skill that needs nurturing. Without close supervision, patients under their care are likely to be mismanaged which would put them at an increased risk of complications and poor prognosis.

Up to 62.7% of participants in the present study believed they were incompetent. This is similar to findings from many previous studies across the world (14–18) but different from some other studies from Sweden(19) and Ireland(20) where most participants believed they were well prepared by medical schools to start their clinical practice. The differences observed in these studies may be explained by variability in teaching methods, availability of resources, and patterns in trainee supervision. Besides, this trend could have changed over time as the population grew and the number of students joining medical schools increased over time since some of these studies were conducted a decade ago. However, it’s worth noting that self-assessment is often influenced by numerous underlying factors such as level of training, prior experiences, and personal biases. For instance, a novice doctor aspiring to become a surgeon may perceive themselves as incompetent simply because they are unable to perform a laparotomy, a procedure that surpasses their current level of training. Considering that intern doctors in Uganda often practice in absence of senior doctors, this could have contributed to the self-percieved incompetence among PRINTs. Among the procedural skills, participants in our study reported having attained reasonable competence in performing basic skills including cannulation, venipuncture, catheterization, and NG tube insertion. This is in agreement with other previous studies assessing competence among fresh graduates and intern doctors(9–11). In Uganda, medical students gain clinical exposure through supervised clinical rotations in year three to five of the five-year medical curriculum, however, the time is quite inadequate for adequate exposure in the different clinical disciplines. Students get opportunities to practice the basic procedural skills during the clinical rottaions albeit for a limited period. Although their training gives limited opportunities if any to learn and practice these skills firsthand, intern doctors in Uganda are required to perform more technical procedures such as Caesarean sections, MVAs, and pleural taps, among others, often times with limited or no supervision. This explains the low self-reported competencies in these procedures. According to our recent study assessing the knowledge and attitude of Basic Life Support (BLS) among undergraduate students in Uganda, it was observed that medical students have limited knowledge of emergency response and care(21). This may explain why many participants in this study believed they were less competent in performing emergency medical procedures. Given the current health system in Uganda where tertiary facilities are mainly the central region, the other regions with lower-level facilities where intern doctors are the main service providers may be less efficient at providing emergency care which contributes to the high mortality rates in the country(22).

Medical students have inadequate time to practice such procedures as lumbar puncture, pleural tap, external drain placements, and hernia repair. Unfortunately, these procedures are increasingly necessary for intern doctors to perform. A novice doctor, who has never performed or even observed a pleural tap is likely to sever the lungs and cause a pneumothorax (23,24). Such knowledge gaps increase the likelihood of inappropriate referrals to tertiary hospitals increasing patient load and limiting potential to provide higher level care to patients who need it (25,26).

One factor hindering the acquisition of surgical skills during training is limited space within the operating theatres amidst growing numbers of trainees. Teaching hospitals have different cadres of health professionals including attending surgeons, fellows, residents, anesthesiology teams, and undergraduate students of different levels. This creates crowding in theatres and denies undergraduate students an ideal environment for learning surgical skills further explaining their skepticism in surgical competencies. Introduction of Virtual Reality in medical training is seen as a high-yielding opportunity to bridge this gap in the following ways; 1) Projection of real-time surgical procedures into a remote spacious room for a comfortable learning environment, and 2) Enabling sharing of video recordings of the performed surgical procedures to students. In settings where virtual reality is still unavailable, skills labs serve as a good alternative where students are given opportunities to practice various basic skills before clinical exposure to improve their confidence and hands-on skills. Although medical schools in Uganda have these facilities, they may be under-utilized.

A good number of participants in this study believed they were competent in OBGY procedures such as; conducting deliveries, C-sections, and MVA’s among others. This is one of the most active departments in medical training where students get direct involvement in patient care. Additionally, due to experiences reported by the intern doctors as regards obstetrics procedures, it becomes prudent for the students in clinical years to look for all possible ways of improving their skills in this discipline. However, majority of participants were still less confident about their levels of competence in this discipline calling for enhanced supervision if these doctors are to perform such procedures in the field.

Although a majority of participants reported having performed or at least observed a majority of the procedures, some of the PRINTs had never witnessed basic procedures like general (venipuncture, NG tube insertion), OBGY (Normal delivery, MVA, and C-section), medical (Paracentesis and Thoracentesis), Surgery (suturing and POP application), and emergency (CPR and airway maneuvers). The probable explanation for this unfortunate observation is the increasing number of students per intake amidst limited resources and mentorship programs. With Uganda standing at a 1:25,000 doctor-patient ratio(27), the demand for doctors is undoubtedly high. However, even with this demand, quality should take precedence over quantity. As the number of trainees increases, resources become limited and learning becomes a competition which puts some trainees at a disadvantage in terms of knowledge and skills. This may explain an increase in the reported cases of medical errors among intern doctors in the country in recent years(4). This effect can be counteracted by mentorship. Mentorship programs prioritize trainee-centered activities that help address their weaknesses and have documented positive impacts on leadership skills, education, biomedical research, and patient care among beneficiaries(28).

In this study, participants with previous work experience in a clinical setting were more likely to report high competence status. This is similar to data from previous studies assessing determinants of clinical competence among healthcare workers(29,30). Work experience in a clinical setting helps boosts one’s confidence to perform soft and procedural skills which explains why participants reported being competent in this assessment. It was observed that the duration of work experience did not determine self-reported competence among our participants. Therefore, measures to improve clinical performance among interns could aim to provide platforms where PRINTs can gain hands-on experience as they wait for their internship program. This would prepare them both in knowledge and provide psychosocial readiness to pursue clinical practice. To the best of our knowledge, this is the first study to assess PRINTs’ readiness to start clinical practice in Uganda. This information could be used by policymakers to address the identified gaps but could also be a stepping stone upon which further inquiries could be based by researchers to determine the reasons underlying the identified gaps.

### Limitations

Our study faced some limitations worth a mention. The response rate was low (65.1%), and it remains unclear if the responses from the participants represent the views of the entire group of current pre-interns in Uganda. Additionally, being a self-administered questionnaire, some of the participants could have misinterpreted some questions and given wrong answers which would cause information bias. However, simple English and guides were used to explain the meaning of the instructions provided for each question to minimize interpretation errors. Using self-assessment as a competence measure poses challenges and can be influenced by other underlying factors, potentially limiting the interpretation and generalizability of our findings. Nevertheless, we firmly believe that a doctor’s self-confidence is vital for effective performance, and any self-doubt should prompt further investigation. Finally, the study assessed only clinical skills and left out other domains of competence which would undermine our conclusions on levels of competence.

## Conclusions

This study shows that majority of PRINTs feel deficient in knowledge and skills to start clinical practice. Although competent in clerkship, participants believed they were less likely to come up with the right diagnoses and management plans.

Participants reported good competence in basic procedural skills such as cannulation but poorest in emergency care. Previous work experience in a clinical setting was the only statistically significant factor associated with self-reported competence. These findings call for urgent interventions bymedical training institutions to improve acquisition of clinical skills by trainees such as increasing supervision and the time alotted to clinical exposure. Skilling programs such as the use of Skills Labs and where possible Virtual Reality need to be integrated into medical training. Pre-internship programs that provide patient exposure experiences such as placements in public health facilities, and Emergence Medical Services to facilitate a smooth transition to clinical practice are recommended. Lastly, further investigation of student learning environments in Ugandan training institutions is necessary to enhance the quality of medical professionals.

## Declarations

### Availability of data and materials

The datasets used and/or analyzed during the current study are available from the corresponding author on reasonable request.

### Authors’ contributions

NS conceptualized the idea. NS, GW, RN, LKS, BS, MN, AN, RA, VNN, SEK, CN and PM developed the study tools with close supervision of PBK. NS, GW, RN, LKS, BS, MN, AN, RA, VNN, SEK, CN and PM collectively participated in the data collection process. NS and GW prepared the first draft of the manuscript and PBK provided extensive comments on the first draft and contributed to developing the final draft. All authors reviewed the final draft and approved the final manuscript.

## Data Availability

All relevant data are within the manuscript and its Supporting Information files.

## Acknowledgment

We commend every doctor who assisted us in shairing our data collection tool. In the same spirit, we appreciate all doctors who spared their time to participate in this study.

## List of abbreviations

CPR: Cardiopulmonary resuscitation
C-Section: Caesarean Section
GCS: Glasgow Coma Scale
GMC: General Medical Council
IUD: Intrauterine device
IV: Intravenous
MBChB: Bachelor of Medicine and Bachelor of Surgery
NG: Nasogastric
OBGY: Obstetrics and Gynecology
POP: Plaster of Paris
PRINTs: Preinterns

## REFERENCES

1. Mwesigwa Alon, Lule Batte Baker, Nakabugo Zurah. Cost of sacking 1,000 doctors. Observer [Internet]. 2017 Nov 20 [cited 2022 Jan 24]; Available from: https://observer.ug/news/headlines/56053-cost-of-sacking-1-000-doctors.html

2. Godfrey O. Poor pay forcing Ugandan doctors to flee to other countries: Health advocates concerned about trend. Anadolu Agency [Internet]. 2021 Nov 26 [cited 2022 Jan 16]; Available from: https://www.aa.com.tr/en/africa/poor-pay-forcing-ugandan-doctors-to-flee-to-other-countries/2431077

3. Alon M. Uganda crippled by medical brain drain | Global development | The Guardian. Global Development [Internet]. 2015 Feb 10 [cited 2022 Jan 16]; Available from: https://www.theguardian.com/global-development/2015/feb/10/uganda-crippled-medical-brain-drain-doctors

4. Wafula P. Daily Monitor. 2020 [cited 2022 Jan 24]. Baby loses arm in botched C-Section operation | Uganda. Available from: https://www.monitor.co.ug/uganda/news/national/baby-loses-arm-in-botched-c-section-operation-1859742?view=htmlamp

5. Dhlamini Zuma. What difference are the many universities in Uganda making? | Uganda. Monitor [Internet]. 2021 Jan 29 [cited 2022 Jan 24]; Available from: https://www.monitor.co.ug/uganda/oped/commentary/what-difference-are-the-many-universities-in-uganda-making--1767336?view=htmlamp

6. Kiguli-Malwadde E, Olapade-Olaopa EO, Kiguli S, Chen C, Sewankambo NK, Ogunniyi AO, et al. Competency-based medical education in two Sub-Saharan African medical schools. Adv Med Educ Pract [Internet]. 2014 Dec 9 [cited 2022 Jan 24];5:483–9. Available from: https://www.dovepress.com/competency-based-medical-education-in-two-sub-saharan-african-medical--peer-reviewed-fulltext-article-AMEP

7. Academy of Medical Royal Colleges. Common Competences Framework for Doctors. Framework. Millbank Media Ltd; 2009.

8. Sample Size Calculator [Internet]. 2023 [cited 2023 Nov 3]. Available from: https://www.calculator.net/sample-size-calculator.html?type=1&cl=95&ci=5&pp=50&ps=500&x=Calculate

9. A P, H D, M H, A G, P M, C E, et al. Self-Perceived Skills of Pre Intern Doctors in Sri Lanka. 2022 Jan 7 [cited 2022 Jan 15]; Available from: https://europepmc.org/article/ppr/ppr440348

10. Obwoge RO, Grave W de. Interns’ Perceived Competency Levels with Respect to the Medical Expert Role in Different Clinical Disciplines. http://www.sciencepublishinggroup.com [Internet]. 2016 Sep 13 [cited 2022 Jan 15];1(1):1. Available from: http://article.sciencepublishinggroup.com/html/10.11648.j.tecs.20160101.11.html

11. Dare A, Fancourt N, Robinson E, Wilkinson T, Bagg W. Training the intern: The value of a pre-intern year in preparing students for practice. 101080/01421590903127669 [Internet]. 2009 [cited 2022 Mar 28];31(8). Available from: https://www.tandfonline.com/doi/abs/10.1080/01421590903127669

12. Busby LP, Courtier JL, Glastonbury CM. Bias in radiology: The how and why of misses and misinterpretations. Radiographics. 2018 Jan 1;38(1):236–47.

13. Bruno MA, Walker EA, Abujudeh HH. Understanding and confronting our mistakes: The epidemiology of error in radiology and strategies for error reduction. Radiographics [Internet]. 2015 Oct 1 [cited 2022 Mar 28];35(6):1668–76. Available from: https://pubs.rsna.org/doi/abs/10.1148/rg.2015150023

14. Ochsmann EB, Zier U, Drexler H, Schmid K. Well prepared for work? Junior doctors’ self-assessment after medical education. BMC Med Educ [Internet]. 2011 [cited 2022 Mar 29];11(1):99. Available from: /pmc/articles/PMC3267657/

15. Morgan PJ, Cleave-Hogg D. Comparison between medical students’ experience, confidence and competence. Med Educ. 2002;36(6):534–9.

16. Muthaura PN, Khamis T, Ahmed M, Hussain SRA. Perceptions of the preparedness of medical graduates for internship responsibilities in district hospitals in Kenya: A qualitative study Curriculum development. BMC Med Educ. 2015 Oct 21;15(1).

17. Jungbauer J, Alfermann D, Kamenik C, Brähler E. Vermittlung psychosozialer kompetenzen mangelhaft: Ergebnisse einer befragung ehemaliger medizinstudierender an sieben deutschen universitäten. PPmP Psychotherapie Psychosomatik Medizinische Psychologie [Internet]. 2003 Jul 1 [cited 2022 Mar 29];53(7):319–21. Available from: http://www.thieme-connect.com/products/ejournals/html/10.1055/s-2003-40493

18. Abuhusain H, Chotirmall S, … NHI medical, 2009 undefined. Prepared for internship? europepmc.org [Internet]. [cited 2022 Mar 29]; Available from: https://europepmc.org/article/med/19489196

19. Hoppe A, Persson E, Birgegárd G. Medical interns’ view of their undergraduate medical education in Uppsala: an alumnus study with clear attitude differences between women and men. Med Teach [Internet]. 2009 [cited 2022 Mar 29];31(5):426–32. Available from: https://pubmed.ncbi.nlm.nih.gov/19811130/

20. Cave J, Woolf K, Jones A, Dacre J. Easing the transition from student to doctor: how can medical schools help prepare their graduates for starting work? Med Teach [Internet]. 2009 [cited 2022 Mar 29];31(5):403–8. Available from: https://pubmed.ncbi.nlm.nih.gov/19142797/

21. Ssewante N, Wekha G, Iradukunda A, Musoke P, Kanyike AM, Nabukeera G, et al. Basic life support, a necessary inclusion in the medical curriculum: a cross-sectional survey of knowledge and attitude in Uganda. BMC Medical Education 2022 22:1 [Internet]. 2022 Mar 3 [cited 2022 Mar 28];22(1):1–8. Available from: https://bmcmededuc.biomedcentral.com/articles/10.1186/s12909-022-03206-z

22. Chamberlain S, Stolz U, Dreifuss B, Nelson SW, Hammerstedt H, Andinda J, et al. Mortality Related to Acute Illness and Injury in Rural Uganda: Task Shifting to Improve Outcomes. PLoS One [Internet]. 2015 Apr 7 [cited 2022 Mar 28];10(4):e0122559. Available from: https://journals.plos.org/plosone/article?id=10.1371/journal.pone.0122559

23. Cantey EP, Walter JM, Corbridge T, Barsuk JH. Complications of thoracentesis: incidence, risk factors, and strategies for prevention. Curr Opin Pulm Med [Internet]. 2016 Jul 1 [cited 2022 Mar 28];22(4):378. Available from: /pmc/articles/PMC8040091/

24. Gordon CE, Feller-Kopman D, Balk EM, Smetana GW. Pneumothorax following thoracentesis: a systematic review and meta-analysis. Arch Intern Med [Internet]. 2010 Feb 22 [cited 2022 Mar 28];170(4):332–9. Available from: https://pubmed.ncbi.nlm.nih.gov/20177035/

25. Yu H, Wang P, Zheng H, Luo J, Liu J. Impacts of congestion on healthcare outcomes: an empirical observation in China. 101080/2327001220201731720 [Internet]. 2020 Jul 2 [cited 2022 Mar 28];7(3):344–66. Available from: https://www.tandfonline.com/doi/abs/10.1080/23270012.2020.1731720

26. Kim SH, Chan CW, Olivares M, Escobar GJ. Association Among ICU Congestion, ICU Admission Decision, and Patient Outcomes. Crit Care Med [Internet]. 2016 Oct 1 [cited 2022 Mar 28];44(10):1814–21. Available from: https://pubmed.ncbi.nlm.nih.gov/27332046/

27. Janepher W. The state of Patient safety in Uganda [Internet]. 2019 [cited 2022 Mar 28]. Available from: https://isqua.org/world-patient-safety-day-blogs/the-state-of-patient-safety-in-uganda.html

28. Choi AMK, Moon JE, Steinecke A, Prescott JE. Developing a culture of mentorship to strengthen academic medical centers. Academic Medicine [Internet]. 2019 May 1 [cited 2022 Mar 28];94(5):630–3. Available from: https://journals.lww.com/academicmedicine/Fulltext/2019/05000/Developing_a_Culture_of_Mentorship_to_Strengthen.25.aspx

29. Taylor I, Bing-Jonsson PC, Finnbakk E, Wangensteen S, Sandvik L, Fagerström L. Development of clinical competence – a longitudinal survey of nurse practitioner students. BMC Nurs [Internet]. 2021 Dec 1 [cited 2022 Mar 28];20(1):1–15. Available from: https://bmcnurs.biomedcentral.com/articles/10.1186/s12912-021-00627-x

30. Takase M. The relationship between the levels of nurses’ competence and the length of their clinical experience: a tentative model for nursing competence development. J Clin Nurs [Internet]. 2013 May [cited 2022 Mar 28];22(9–10):1400–10. Available from: https://pubmed.ncbi.nlm.nih.gov/22957733/

